# DNA methylation and gene expression pattern of *ACE2* and *TMPRSS2* genes in saliva samples of patients with SARS-CoV-2 infection

**DOI:** 10.1101/2020.10.24.20218727

**Authors:** Pratibha Misra, Bhasker Mukherjee, Rakhi Negi, Vikas Marwah, Arijit Kumar Ghosh, Prashant Jindamwar, Mukesh U Singh, Y Vashum, R Syamraj, G Bala Chandra, M K Sibin

**Affiliations:** Department of Biochemistry, Armed Forces Medical College, Pune, India; Department of Respiratory Medicine, Army Institute of Cardio thoracic sciences, Pune, India; Department of Cardiology, Army Institute of Cardio thoracic sciences, Pune, India; Department of Lab Sciences, Army Institute of Cardio Thoracic Sciences, Pune, India

**Keywords:** COVID-19, ACE2 and TMPRSS2, Non-invasive sample, Epigenetic modification

## Abstract

COVID-19 caused by SARS-CoV-2 became a pandemic affecting the health and economy of the world. Although it was known that this virus uses *ACE2* protein along with *TMPRSS2* to enter the host cell, the methylation pattern and gene expression of *ACE2* and *TMPRSS2* genes are not explored in saliva samples of patients infected with COVID-19. The study aimed to quantify promoter methylation of *ACE2* and *TMPRSS2* along with its mRNA expression in saliva samples of COVID-19 patients in order to understand the regulatory mechanism of these genes in SARS-CoV-2 infection. Saliva samples were collected from thirty male patients with SARS-CoV-2 infection and thirty age-matched healthy control male subjects. Q MS PCR and qRT PCR was performed to quantify the promoter DNA methylation and mRNA expression of *ACE2* and *TMPRSS2* respectively. Our study didn’t find any significant difference between methylation and expression of these two genes in cases compared to control subjects. However there was significant positive correlation between DNA methylation of *ACE2* and its gene expression. Among cases, the sample collected ≥7 days after appearance of symptoms showed higher amount of methylation in both *ACE2* and *TMPRSS2* genes when compared to sample collected before 7 days. In conclusion, we found that *ACE2* and *TMPRSS2* methylation plays a role in COVID-19.

## 1. Introduction

Coronavirus disease 2019 (COVID-19), leading to clusters of severe respiratory illness, became a pandemic with a total of 39 million reported positive cases as of 16 October 2020 [World Health Organization, 2020]. It is caused by severe acute respiratory syndrome coronavirus 2 (SARS-CoV-2) which is a member of Beta-CoV lineage B [Zhu et al, 2019]. It is positive stranded RNA viruses with a protein envelope around it. Protein envelop contains spike glycoprotein with two sub units S1 and S2. The S1 is involved in the attachment of virus to host cell membrane [Belouzard et al, 2012]. Recently, it is reported that virus uses ACE2 (Angiotensin converting enzyme-2) protein along with Transmembrane Protease Serine 2 (TMPRSS2) to enter the host cell [Kupferschmidt and Cohen, 2020].

ACE2 is an enzyme found in the cell membrane of various tissues and is involved in counterbalancing ACE. ACE2 is present on cell membrane of various human tissues like lung, liver, pancreas and blood. It was reported as the most important receptor of SARS-CoV-2 in lung epithelial cells [Zill et al, 2012, Rui and Sang, 2020]. The gene expression of *ACE2* in cell is controlled epigenetically [Zill et al, 2012]. The promoter methylation of this gene is lowest among lung epithelial cells and thus expression rates are high in this tissue [Corley and Ndhlovu, 2020]. It was also observed that methylation of *ACE2* gene is important in COVID-19 and it also correlated with age and gender of individuals [Corley and Ndhlovu, 2020, Pinto et al, 2020]. Previous study conducted on 700 lung transcriptome samples showed that there was over expression of *ACE2* in lung tissue [Pinto et al, 2020]. *ACE2* gene is generally hypermethylated in respiratory system of children [Corley and Ndhlovu, 2020] and this explains why older individuals are more susceptible to SARS-CoV-2 when compared to children.

TMPRSS2 is a protein found in various tissues. TMPRSS2 is an Androgen Receptor signalling downstream gene which is closely related to prostate carcinogenesis. DNA methylation is a key mechanism to influence gene expression in *TMPRSS2* [Chu et al, 2014]. Protein expression studies showed that TMPRSS2 is highly expressed in lung epithelial cells when compared to other cells in lungs [Bertram et al, 2012]. It was reported that TMPRSS2 is required for entry and spread of SARS-CoV-2 in human [Hoffmann et al, 2020, Matsuyama et al, 2010]. During SARS-CoV-2 infection, TMPRSS2 causes priming of viral spike glycoprotein by cleaving it and help in viral entry through fusion of viral and cellular membrane [Hoffmann et al, 2020].

However, the methylation pattern and gene expression of *ACE2* and *TMPRSS2* genes are not explored in saliva samples of patients infected with COVID-19. The correlation of methylation status with gene expression of these genes gives a comprehensive idea about role of these genes in pathogenesis of COVID-19 infection. Present study aimed to quantify promoter methylation of *ACE2* and *TMPRSS2* along with its mRNA expression in saliva samples of COVID-19 patients in order to understand the regulatory mechanism of these genes in SARS-CoV-2 infection.

## 2. Methodology

### Recruitment of subjects

This case-control study was conducted at department of biochemistry, Armed forces medical college, Pune (AFMC) and COVID-19 ward of Army institute of cardio thoracic sciences (AICTS), Pune, over a period of two months (August-September 2020). Ethical clearance was taken from the Institutional Ethics Committee, AFMC (S No. IEC/2020/95). After obtaining written informed consent, 30 random subjects who were tested positive for SARS-CoV-2 infection by a reverse transcription-polymerase chain reaction (RT-PCR) were recruited in the study. All patient samples were collected from the COVID-19 ward where patients were admitted as per national and Indian Armed Forces guidelines. Patients who were below 18 years of age, above 60 years and patients with severe illness were excluded from the study. Thirty age matched individuals who were asymptomatic, tested negative using antibody test for SARS-CoV-2 and without any comorbidities were included as healthy control subjects in the study.

### Sample collection

Saliva samples were collected as per Centers for Disease Control and Prevention (CDC) guidelines from all subjects by a trained healthcare professional. All subjects were provided with a wide mouthed sterile container and asked to spit the saliva into the container. The collection containers were sealed inside the COVID-19 ward using parafilm and kept in an outer plastic container which was then packed in sample collection box which maintain a temperature of 18-20°C during transport. Samples were processed for DNA and RNA extraction in a Biosafety level-3 (BSL-3) laboratory within 3 hours of sample collection.

### Nucleic acid extraction

Total RNA was isolated using High Pure RNA isolation kit (Roche diagnostics) following the manufacturer’s instructions. DNA was extracted, using DNeasy Blood and tissue kit (Qiagen) according to manufacturer’s protocol. DNA/RNA was quantified using Nanodrop spectrophotometer, NanoDrop ND 2000c (Thermo). DNA samples having 1.75–1.90 (A_260/280_) and RNA samples having 1.95–2.05 (A_260/280_) were included for further analysis.

### Bisulfite conversion and qMS (quantitative methylation specific) PCR

DNA samples were bisulfite modified using EpiTect Fast Bisulfite kit (Qiagen, Germany) as per manufacturer’s protocol. Analysis of *ACE2* and *TMPRSS2* promoter methylation was performed using real time PCR with TB Green Premix Ex Taq II mastermix (Takara). β-*actin (ACTB)* unmethylated primers were used to amplify the beta actin gene which was used as control for DNA input. PCR was performed on Quantstudio 5 instrument (Applied Biosystems) as per the temperature and conditions provided with mastermix. Universal poly methylated control DNA was used as positive control and nuclease free water was used as no template control. Cycle threshold (Ct) or Cycle quantitation (Cq) values were collected for *ACE2, TMPRSS2* and *ACTB*. Amount of *ACE2, TMPRSS2* promoter methylation was determined by 2^(−ΔCt) comparative analysis method [Schmittgen et al, 2008]. Melt curve analysis was performed with each set of primers to check the non-specific amplification or primer dimer formation. Primers were designed using Gene Runner software (Version 3.05). Details of primers used are given in Table 1. Locations of CpG island and primer binding is shown in Figure 1.

**Table 1.**
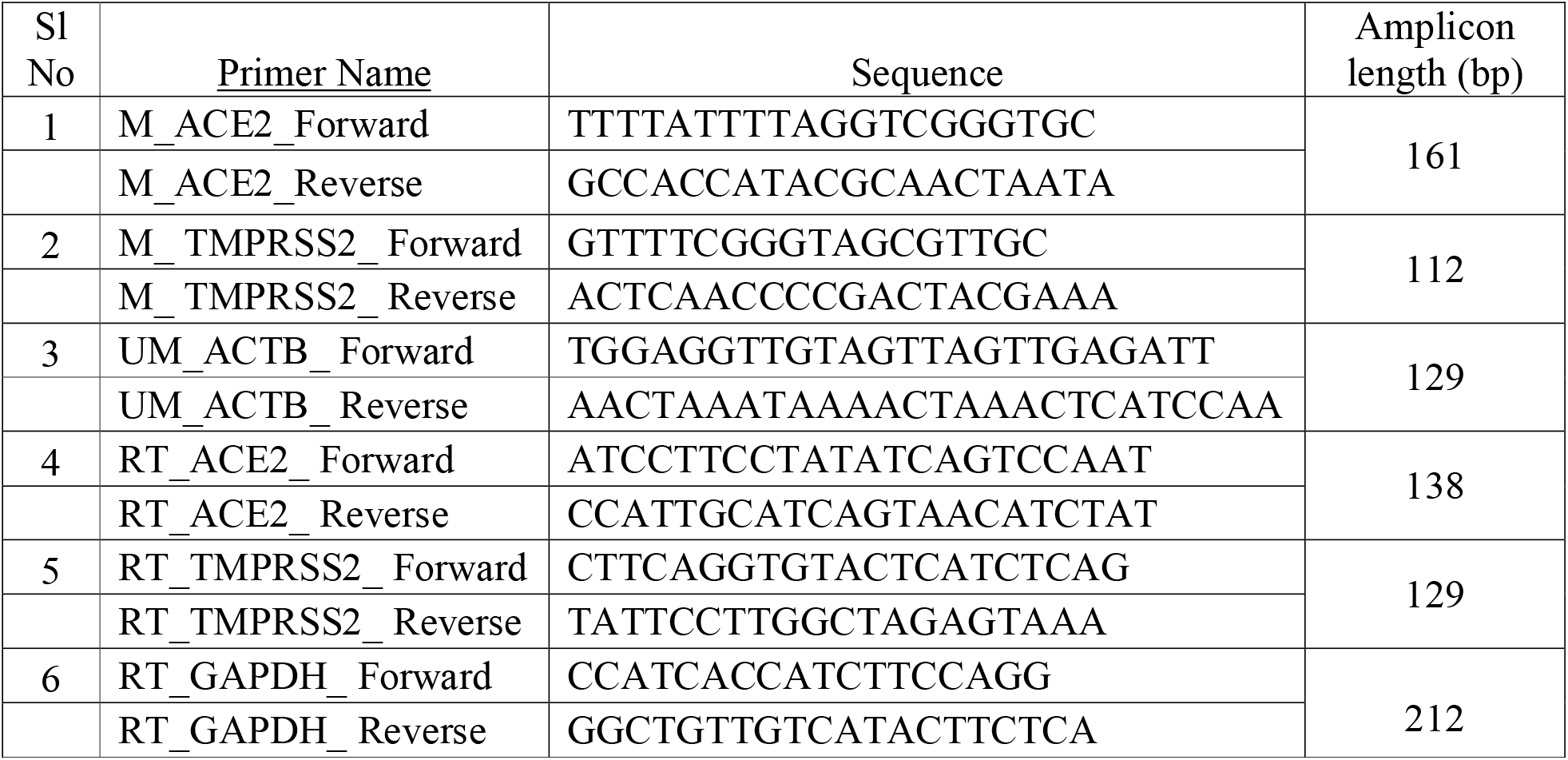
Details of primers used in qMS-PCR and qRT-PCR

**Figure 1:**
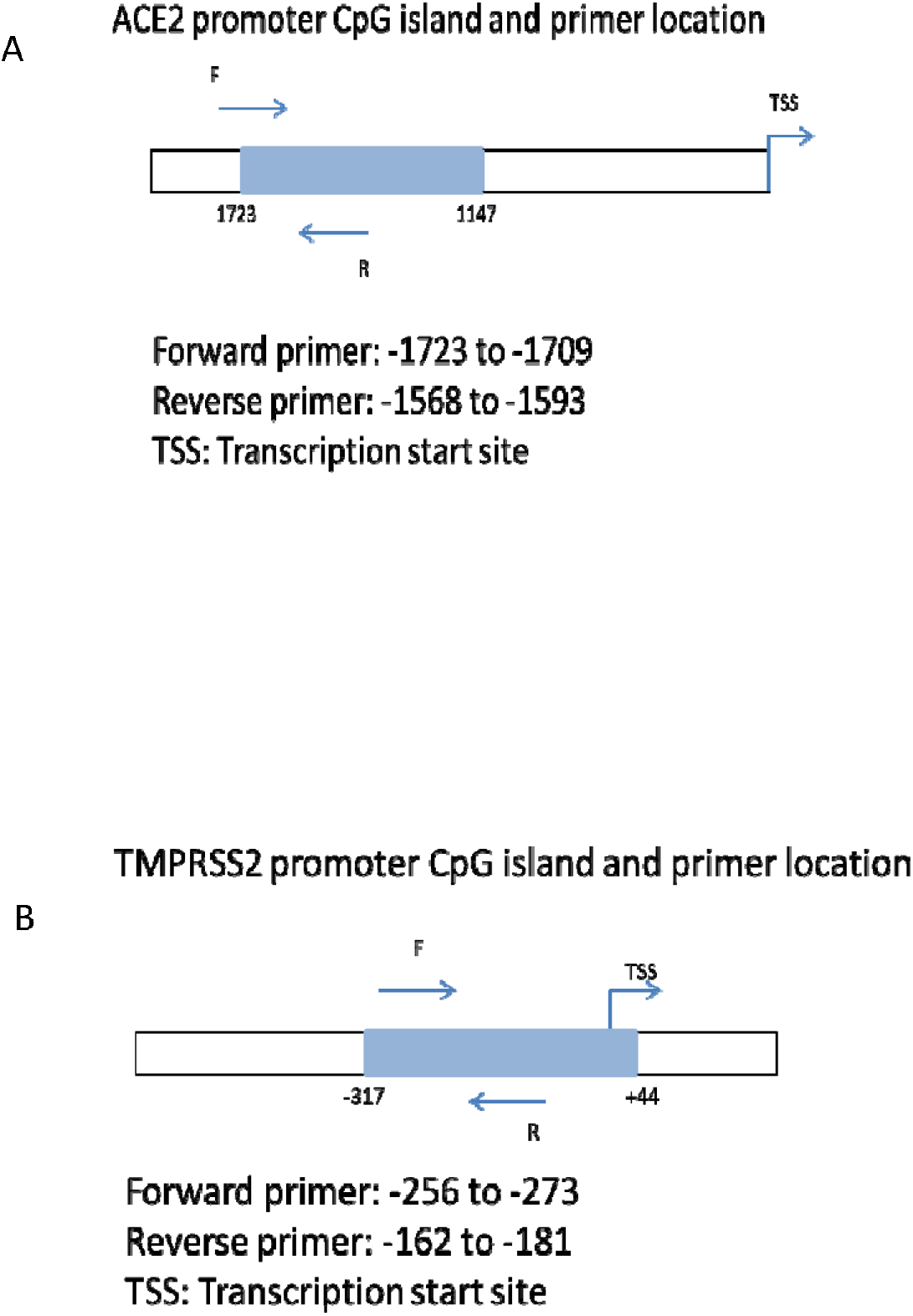
Promoter region with CpG island and primer sequence of (A) *ACE2* gene and (B) *TMPRSS2* gene

### Reverse Transcription and qPCR

500 Nano grams of total RNA was reverse transcribed into complementary DNA (cDNA) using PrimeScript 1^st^ strand cDNA synthesis kit (Takara) following manufacturer’s directions. *ACE2* and *TMPRSS2* mRNA expression was quantified using TB Green Premix Ex Taq II (Takara) in duplicates. Glyceraldehyde 3-phosphate dehydrogenase (GAPDH) was used as housekeeping gene to normalise the expression. Comparative expression analysis by 2^(−ΔCt) method was used to compare the expressions. Sequences of the primers are given in Table 1.

### Statistical analysis

The data were analysed using R 3.5.2 version software for Windows. Categorical variables were given as counts and numerical variables were represented as median. Shapiro Wilk test was used for normality testing of the continuous variables. As the data was not normally distributed they were analysed using non-parametric tests. Wilcoxon two sample test was used to compare two groups. Spearman correlation test was performed to compare methylation and gene expression data. The χ2 test was used to check association between categorical variables. P value less than 0.05 was considered significant.

## 3. Results

The continuous variables in the study data were not normally distributed therefore the values were represented as median. The median age of the patient group was 31.0 years which was not significantly different from healthy control median age of 32.5 years (p=0.116). Eighty per cent of patients were less than 40 years of age and 60 per cent were symptomatic at the time of study. The demographic characteristics of the patients are summarized in the Table 2.

**Table 2.** Demographic characteristics of the patients and controls

| Variable | Cases |  | Control | P value |
| --- | --- | --- | --- | --- |
| Age (in years-median) | 31.0 |  | 32.5 | 0.116 |
| Age group | <40 years | 24 | 21 | 0.371 |
|  | ≥40 years | 6 | 9 |  |
| Symptoms | Symptomatic | 18 | - | - |
|  | Asymptomatic | 12 | - |  |
| Co morbidity | Yes<br>(Hypertension) | 5 | - |  |
|  | No | 25 | - |  |
| Clinical presentation | Mild | 27 | - |  |
|  | Moderate | 3 | - |  |
| Day of sample collection | <7 days | 10 | - |  |
|  | ≥7 days | 20 | - |  |

### *ACE2* and *TMPRSS2* methylation and expression in cases and controls

Gene expression data of all 30 cases and controls were available for analysis, but due to quality issues in DNA, only 21 cases and 20 control data of methylation for *ACE2* and *TMPRSS2* were available. There was no significance difference between *ACE2* methylation in cases and controls (p=0.294). Similarly, there was no significance difference between *ACE2* expression in cases and controls (p=0.929). There was no significance difference between *TMPRSS2* methyaltion in cases and controls (p=0.498). Likewise there was no significance difference between *TMPRSS2* expression in cases and controls (p=0.559) (Table 3, Figure 2).

**Table 3.**
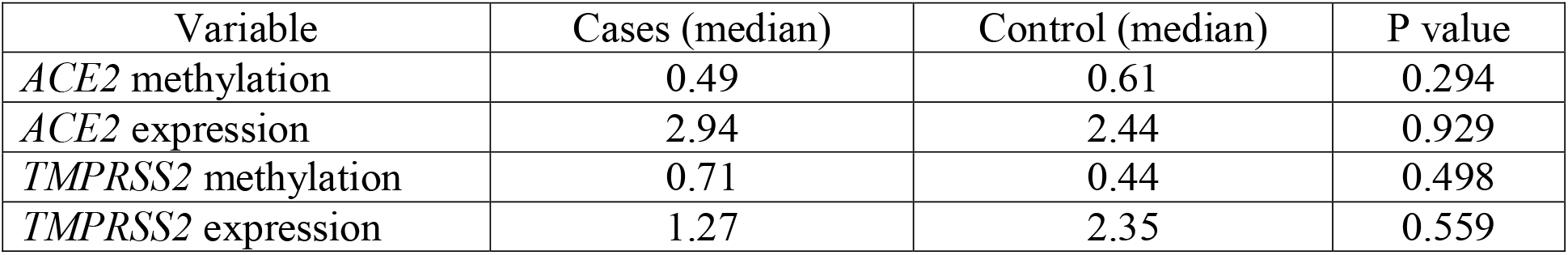
*ACE2/TMPRSS2* methylation/expression data of the patients and controls

**Figure 2:**
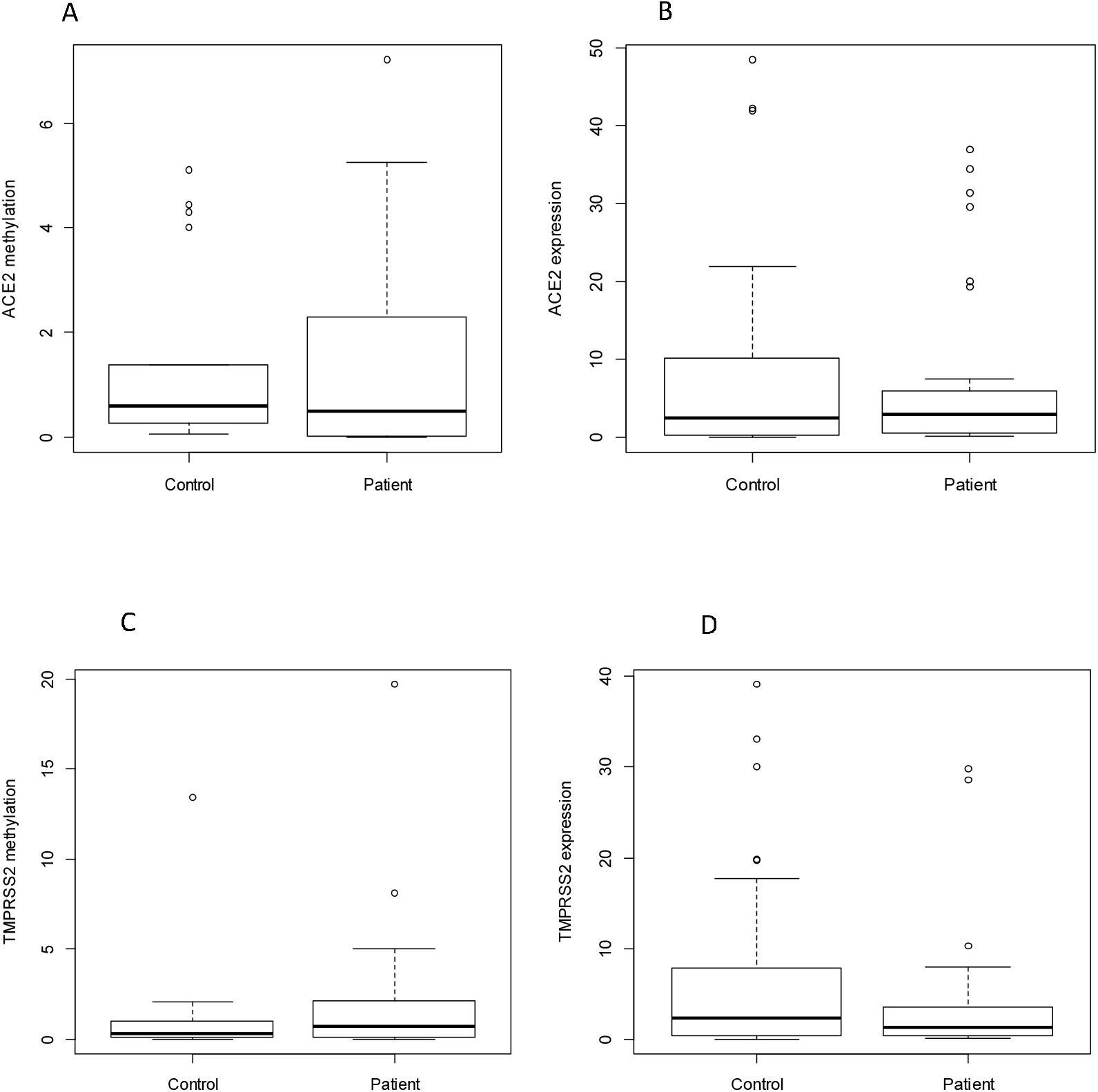
(A) *ACE2* methylation in cases and controls, (B) *ACE2* expression in cases and controls, (C) *TMPRSS2* methylation in cases and controls, (D) *TMPRSS2* expression in cases and controls

### Correlation of *ACE2* and *TMPRSS2* methylation and gene expression

There was a significant negative correlation between *ACE2* expression and *ACE2* methylation (rho= -0.342, p value= 0.031) but the correlation between *TMPRSS2* expression and *TMPRSS2* methylation was not significant (rho= -0.251, p value= 0.118). There was significant positive correlation between *ACE2* expression and *TMPRSS2* expression (rho= 0.332, p value= 0.009). But there was no significant correlation between *ACE2* methylation and *TMPRSS2* methylation (rho= 0.307, p value= 0.054) (Figure 3).

**Figure 3:**
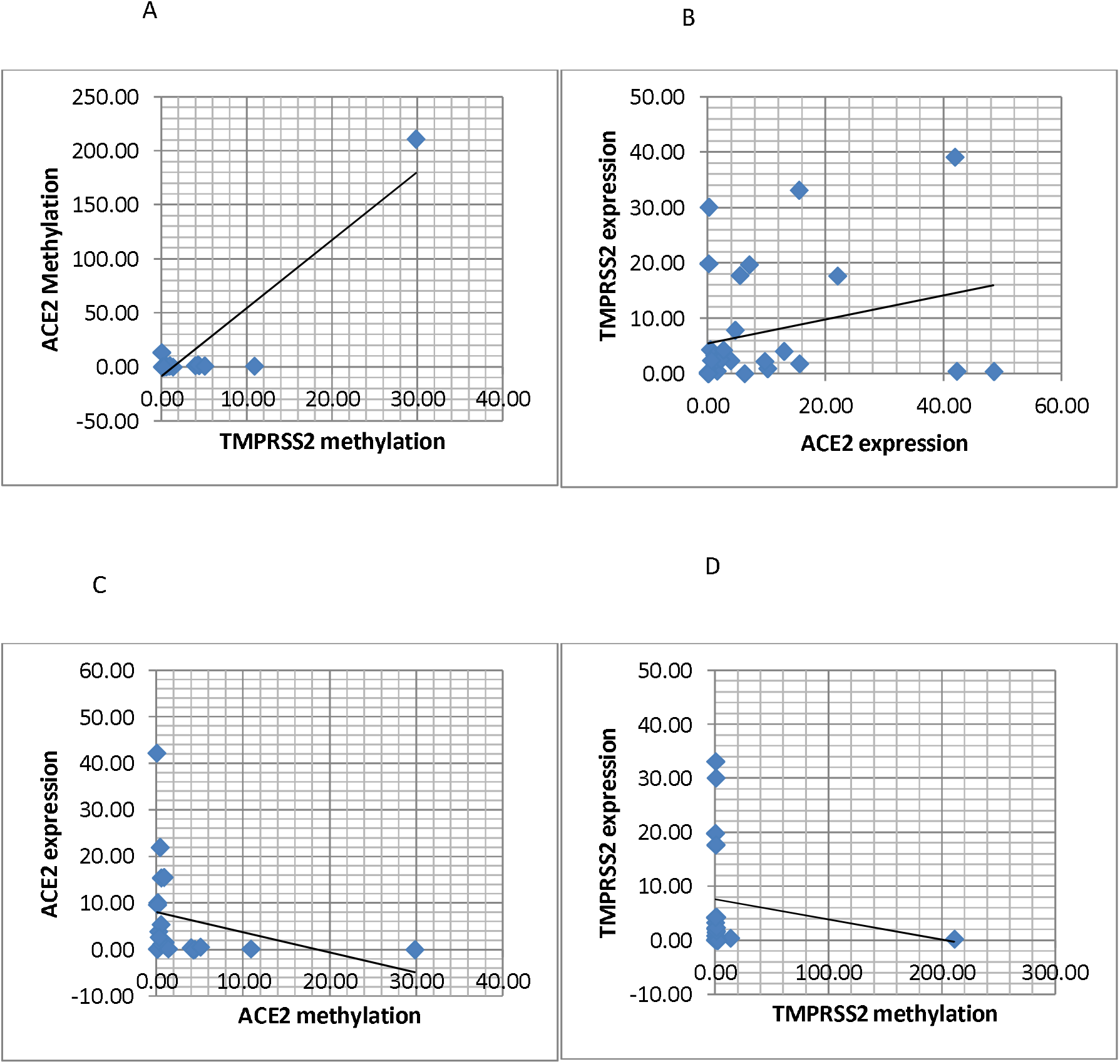
Correlation of (A) *ACE2* methylation with *TMPRSS2* methylation, (B) *ACE2* expression with *TMPRSS2* expression, (C) *ACE2* methylation with *ACE2* expression, (D) *TMPRSS2* methylation with *TMPRSS2* expression

### *ACE2* and *TMPRSS2* methylation and expression with clinical correlation

The methylation of *ACE2* and *TMPRSS2* genes significantly differed in relation to the day of saliva sample collection.(Figure 4). The samples collected more than or equal to 7 days after appearance of symptoms showed higher amount of methylation in *ACE2* (p=0.009) and *TMPRSS2* (0.042). There was no association between clinical variables and expression or methylation of both genes. Data is represented in Table 4.

**Table 4.** Association of *ACE2* and *TMPRSS2* methylation and expression with clinical variables

| Variable | *ACE2*<br>methylation | P<br>value | *ACE2*<br>expression | P<br>value | *TMPRSS2*<br>methylation | P<br>value | *TMPRSS2*<br>expression | P<br>value |
| --- | --- | --- | --- | --- | --- | --- | --- | --- |
| Age group |  |  |  |  |  |  |  |  |
| <40 years | 0.49 | 0.698 | 2.37 | 0.743 | 0.60 | 0.687 | 1.27 | 0.876 |
| ≥40 years | 1.54 |  | 3.21 |  | 1.39 |  | 1.51 |  |
| Symptoms |  |  |  |  |  |  |  |  |
| Symptoma<br>tic | 0.36 | 0.382 | 2.69 | 0.983 | 1.16 | 0.670 | 1.04 | 0.916 |
| Asympto<br>matic | 1.84 |  | 2.94 |  | 0.37 |  | 1.43 |  |
| Co morbidity |  |  |  |  |  |  |  |  |
| Yes | 3.03 | 0.412 | 2.66 | 0.589 | 1.73 | 0.451 | 1.08 | 0.718 |
| No | 0.39 |  | 3.23 |  | 0.60 |  | 1.41 |  |
| Day of sample collection |  |  |  |  |  |  |  |  |
| <7 days | 0.02 | **0.009** | 2.17 | 1 | 0.12 | **0.042** | 1.68 | 0.281 |
| ≥7 days | 1.50 |  | 3.21 |  | 1.50 |  | 1.10 |  |
| Clinical presentation |  |  |  |  |  |  |  |  |
| Mild | 0.49 | 0.857 | 2.66 | 0.845 | 0.60 | 0.590 | 1.41 | 1 |
| Moderate | 3.61 |  | 3.76 |  | 1.49 |  | 1.08 |  |

**Figure 4:**
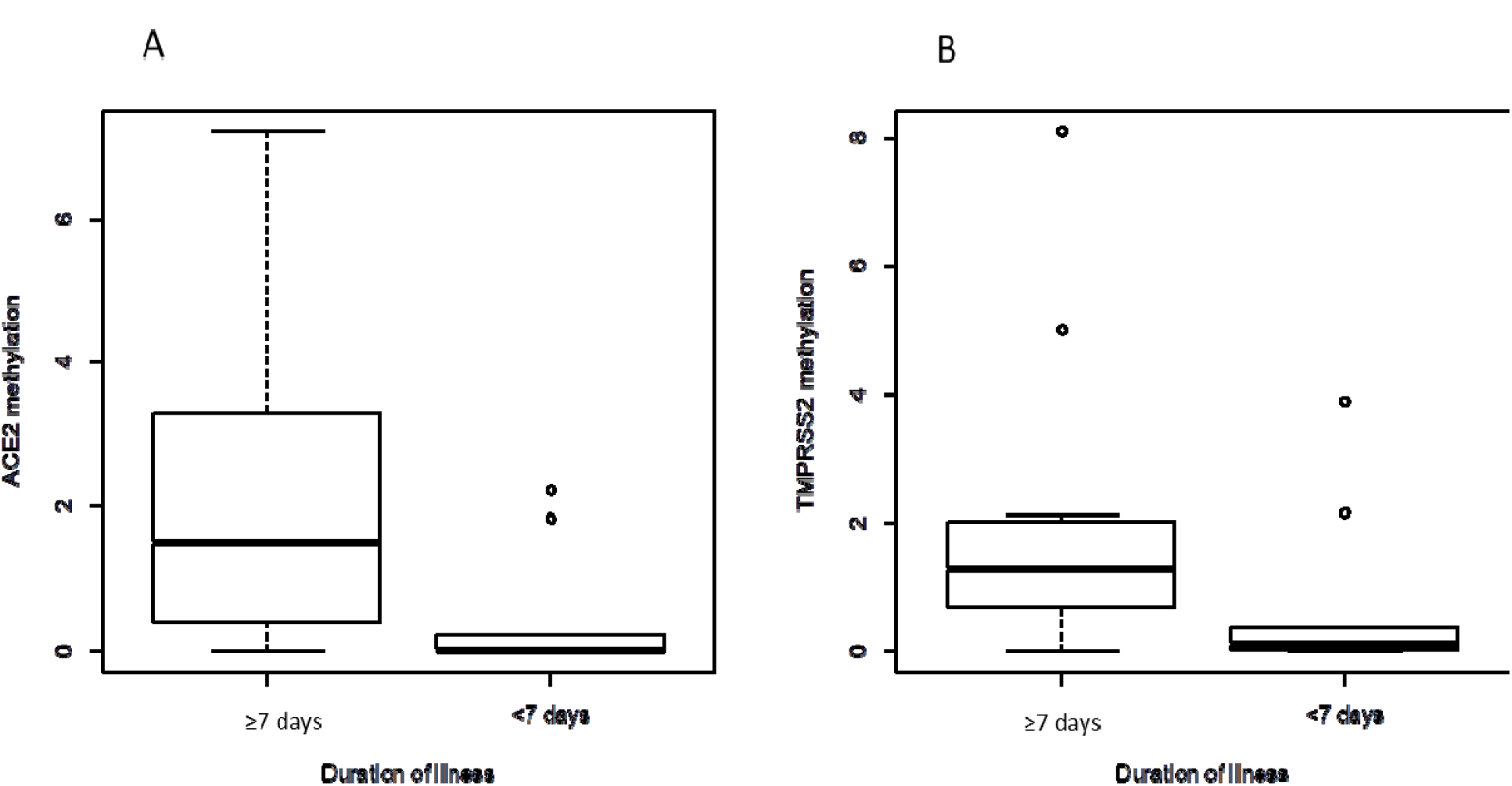
Association of (A) *ACE2* methylation in patients with <7 days of illness and ≥7 days of illness, (B) *TMPRSS2* methylation in patients with <7 days of illness and ≥7 days of illness

## 4. Discussion

Although it is known that ACE2 and TMPRSS2 proteins are involved as receptors in SARS-CoV-2 entry into host cell, regulatory mechanism of its expression in host cell after the SARS-CoV2 infection is not well known. Upon the entry of a virus into host cell, activation of various molecular signalling pathways may occur which help the virus to proliferate and transmit inside the host [Yogendra et al, 2020]. Epigenetic modifications are also reported in host cell DNA after the entry and proliferation of the virus [Liu et al, 2019]. In the present study we tried to decipher the epigenetic regulation mediated mRNA expression of *ACE2* and *TMPRSS2* genes in saliva samples of patients infected with SARS-CoV2 in comparison with healthy control subjects. Our study was a preliminary attempt and first of its kind to report the *ACE2* and *TMPRSS2* gene methylation and expression in saliva samples of SARS-CoV2 infected individuals.

*ACE2* mRNA and protein was reported to be abundantly expressed in epithelial cells of oral mucosa and saliva samples [Srinivasan et al, 2020]. The *TMPRSS2* expression was also found in saliva samples but it largely varied from undetectable to very high concentration in individual samples [Xu et al, 2020]. But two studied mentioned above were conducted on non COVID 19 samples. So measuring *ACE2* and *TMPRSS2* expression in saliva sample in COVID-19 cases could be a way to assess the susceptibility to SARS-CoV-2 infection when compared to invasive nasopharyngeal or lung tissue. We carried out expression study of *ACE2* and *TMPRSS2* in RNA isolated from saliva samples of SARS-CoV2 infected cases and healthy controls to see any changes in expression due to infection. However, the expression level in cases was not significantly different from controls. The expressions of these genes were also not associated with any of the clinical variable studied. Pinto et al, showed that ACE2 expression is upregulated in lungs of COVID-19 patients with co-morbidities and severity [Pinto et al, 2020]. But our study included only mild and moderate cases and very few numbers of cases with co morbidities. This may be the reason for not getting significant differences in cases and controls. The expression of *ACE2* in nasal epithelium is shown to change with age, lowest in younger children, high in adolescents and highest in adults [Bunyavanich et al, 2020]. But in our study we didn’t get any significant difference in age below 40 years and above 40 years. It may be because we included only subjects in the age group 18-60, which include mainly adults who were young and middle aged. The association of *ACE2* expression with the gender was inconsistent as per available literature [Xie et al, 2006; Soro-Paavonen et al, 2012, Dalpiaz et al, 2015]. But testosterone and estrogen were known to influence *ACE2* expression (Glinsky, 2020; Li et al, 2020]. Even *TMPRSS2* expression was controlled by androgen hormones as it is an androgen regulated protein [Shirogane et al, 2008]. In order to avoid this confounding we had included only male subjects in the study.

*ACE2* and *TMPRSS2* expression is controlled by various regulatory mechanisms in the cell. [FitzGerald et al, 2008; Asselta et al, 2020; Zill et al, 2012; Chu et al, 2014]. In an Italian cohort, influence of genetic variants of *ACE2* and *TMPRSS2* on its expression was observed [FitzGerald et al, 2008; Asselta et al, 2020]. These genes are also shown to be regulated by epigenetics [Zill et al, 2012; Chu et al, 2014]. It was predicted that there is differential methylation in *ACE2* gene and that may be the cause of variability of its expression and it could be used to check the susceptibility of an individual to SARS-CoV-2 infection [Steyaert et al, 2020]. However, promoter DNA methylation of both *ACE2* and *TMPRSS2* was not quantified in subjects infected with SARS-CoV-2. In our study, the methylation level of *ACE2* and *TMPRSS2* in cases was not significantly different from controls. This could be due to small sample size comprising of mild to moderate cases.

During analysis we found that both *ACE2* and *TMPRSS2* showed the classic pattern of methylation inversely proportional to expression in all samples. The saliva samples were taken at single point in time from cases. The quantitative methylation level however did reveal difference between samples from patients drawn less than 7 days of symptoms and ≥7 days post symptomatic. The methylation of both *ACE2* and *TMPRSS2* were higher in cases who were ≥7 day post symptomatic. This indicates that promoter methylation modification occurs after few days of infection and it may lead to reduced expression of these genes post infection. In mouse lung tissue, *ACE2* expression was reduced to a considerable amount after SARS-CoV infection showing the down-regulation of *ACE2* expression post infection [Kuba et al, 2005]. Even H1N1 influenza infection showed down-regulation of *ACE2* [Liu et al, 2014]. Both these studies support our results. We also hypothesise that mild cases may show higher methylation in these genes leading to low expression as a host defence and in severe cases with co-morbidities the methylation may be reduced to have high expression and leading to catastrophic consequences among this group. In our study, the methylation and expression of *ACE2* showed positive correlation with that of *TMPRSS2*. This finding emphasizes on the coordination of both *ACE2* and *TMPRSS2* proteins in acquiring SARS-CoV2 infection.

Our study has some strength points as we have included only male genders to avoid the confounding of sex hormones. We have excluded individuals below 18 years and above 60 years of age to avoid confounding with the methylation/expression levels. Clinical samples used were non-invasive in nature which is an advantage. We have also performed both methylation and mRNA experiments together so that the data is consistent. The RNA and DNA samples extracted were of good quality and quantity. Limitations of our study include small sample size and exclusion of severe cases. Saliva samples may contain white blood cells which may contribute DNA that can be a confounder [Theda et al, 2018]. In future, a follow up study with patients with SARS-CoV-2 during infection and after recovery will give more insight on host epigenetic adaptation. A study with mild/moderate versus severe cases may delineate the pathological role of *ACE2*/ *TMPRSS2* expression in severe cases.

## 5. Conclusion

We report that *ACE2* and *TMPRSS2* methylation and expression in saliva samples were not significantly different in SARS-CoV-2 positive subjects when compared to healthy controls. However, *ACE2* methylation correlate negatively with its expression and *ACE2* expression was positively correlated with *TMPRSS2* expression. Methylation of *ACE2* and *TMPRSS2* was significantly higher in patients with SARS-CoV2 infection from whom the sample was collected ≥7 day after appearance of symptoms, which suggests that there is epigenetics modification in the host after a particular period of initial infection. Further studies with large sample size including severe cases with co morbidities may give more information about epigenetic modification of these two genes in SARS-CoV2.

## Data Availability

All raw data will be available on request

## DECLARATIONS

Ethics approval and consent to participate: Ethical clearance was taken from the Institutional Ethics Committee, AFMC (S No.IEC/2020/95). Written informed consent was taken from all participants enrolled in the study.

## Authors contribution

**Pratibha Misra:** Conceptualization and designing methodology of the work, analysis and interpretation of data, writing original draft, critical revision of the article & final approval of the version to be published. **Bhasker Mukherjee:** data curation and editing of the article. **Rakhi Negi:** data curation and editing of the article. **Vikas Marwah:** Investigation and acquisition of data. **Arijit Kumar Ghosh:** Investigation and acquisition of data. **Prashant Jindamwar:** Investigation and acquisition of data. **Mukesh U:** Acquisition of data **and a**nalysis. **Vashum Y:** Data analysis. **Shyamraj R:** Investigation. **G Bala Chandra:** Investigation. **Sibin M K:** Methodology of work, acquisition, analysis and interpretation of data, writing original draft and final revision of the article. All authors read and approved the final manuscript.

## Acknowledgements

We acknowledge Lt Gen Nardeep Naithani, Director & Commandant, Armed Forces Medical College, Pune & Maj Gen Arindam Chatterjee, Commandant, Army Institute of Cardio Thoracic Sciences, Pune for their guidance and kind support.

## References

1. Asselta, R., E. M. Paraboschi, A. Mantovani and S. Duga, 2020 ACE2 and TMPRSS2 variants and expression as candidates to sex and country differences in COVID-19 severity in Italy.

2. Belouzard, S., J. K. Millet, B. N. Licitra and G. R. Whittaker, 2012Mechanisms of coronavirus cell entry mediated by the viral spike protein. Viruses 4: 1011–1033.

3. Bertram, S., A. Heurich, H. Lavender, S. Gierer, S. Danisch et al., 2012Influenza and SARS-coronavirus activating proteases TMPRSS2 and HAT are expressed at multiple sites in human respiratory and gastrointestinal tracts. PloS one 7: e35876.

4. Bunyavanich, S., A. Do and A. Vicencio, 2020Nasal gene expression of angiotensin-converting enzyme 2 in children and adults. Jama.

5. Cai, H., 2020Sex difference and smoking predisposition in patients with COVID-19. The Lancet Respiratory Medicine 8: e20.

6. Chu, M., Y. Chang, N. Wang, W. Li, P. Li et al., 2014Hypermethylation-mediated transcriptional repression of TMPRSS2 in androgen receptor-negative prostate cancer cells. Experimental Biology and Medicine 239: 823–828.

7. Corley, M. J., and L. C. Ndhlovu, 2020 DNA methylation analysis of the COVID-19 host cell receptor, angiotensin I converting enzyme 2 gene (ACE2) in the respiratory system reveal age and gender differences.

8. Dalpiaz, P., A. Lamas, I. Caliman, R. Ribeiro Jr, G. Abreu et al., 2015Sex hormones promote opposite effects on ACE and ACE2 activity, hypertrophy and cardiac contractility in spontaneously hypertensive rats. PLoS One 10: e0127515.

9. FitzGerald, L. M., I. Agalliu, K. Johnson, M. A. Miller, E. M. Kwon et al., 2008Association of TMPRSS2-ERG gene fusion with clinical characteristics and outcomes: results from a population-based study of prostate cancer. BMC cancer 8: 230.

10. Glinsky, G., 2020 Genomics-guided molecular maps of coronavirus targets in human cells: a path toward the repurposing of existing drugs to mitigate the pandemic. arXiv preprint 2003.13665.

11. Hoffmann, M., H. Kleine-Weber, S. Schroeder, N. Krüger, T. Herrler et al., 2020SARS-CoV-2 cell entry depends on ACE2 and TMPRSS2 and is blocked by a clinically proven protease inhibitor. Cell.

12. Kuba, K., Y. Imai, S. Rao, H. Gao, F. Guo et al., 2005A crucial role of angiotensin converting enzyme 2 (ACE2) in SARS coronavirus–induced lung injury. Nature medicine 11: 875–879.

13. Kupferschmidt, K., and J. Cohen, 2020Race to find COVID-19 treatments accelerates, pp. American Association for the Advancement of Science.

14. Leung, J. M., C. X. Yang, A. Tam, T. Shaipanich, T.-L. Hackett et al., 2020ACE-2 expression in the small airway epithelia of smokers and COPD patients: implications for COVID-19. European Respiratory Journal 55.

15. Li, R., S. Qiao and G. Zhang, 2020Analysis of angiotensin-converting enzyme 2 (ACE2) from different species sheds some light on cross-species receptor usage of a novel coronavirus 2019-nCoV. Journal of Infection 80: 469–496.

16. Li, Y., W. Zhou, L. Yang and R. You, 2020Physiological and pathological regulation of ACE2, the SARS-CoV-2 receptor. Pharmacological research: 104833.

17. Liu, S., L. Liu, G. Xu, Z. Cao, Q. Wang et al., 2019Epigenetic modification is regulated by the interaction of influenza A virus nonstructural protein 1 with the de novo DNA methyltransferase DNMT3B and subsequent transport to the cytoplasm for K48-linked polyubiquitination. Journal of virology 93.

18. Liu, X., N. Yang, J. Tang, S. Liu, D. Luo et al., 2014Downregulation of angiotensin-converting enzyme 2 by the neuraminidase protein of influenza A (H1N1) virus. Virus research 185: 64–71.

19. Matsuyama, S., N. Nagata, K. Shirato, M. Kawase, M. Takeda et al., 2010Efficient activation of the severe acute respiratory syndrome coronavirus spike protein by the transmembrane protease TMPRSS2. Journal of virology 84: 12658–12664

20. Organization, W. H., and W. h. organization, 2020 Coronavirus disease (COVID-2019) situation reports, pp

21. Pinto, B. G., A. E. Oliveira, Y. Singh, L. Jimenez, A. N. A. Gonçalves et al., 2020ACE2 expression is increased in the lungs of patients with comorbidities associated with severe COVID-19. MedRxiv

22. Schmittgen, T. D., and K. J. Livak, 2008Analyzing real-time PCR data by the comparative C T method. Nature protocols 3: 1101

23. Shirogane, Y., M. Takeda, M. Iwasaki, N. Ishiguro, H. Takeuchi et al., 2008Efficient multiplication of human metapneumovirus in Vero cells expressing the transmembrane serine protease TMPRSS2. Journal of virology 82: 8942–8946

24. Singh, Y., G. Gupta, S. Satija, K. Pabreja, D. K. Chellappan et al., 2020COVID[19 transmission through host cell directed network of GPCR. Drug Development Research

25. Soro-Paavonen, A., D. Gordin, C. Forsblom, M. Rosengard-Barlund, J. Waden et al., 2012Circulating ACE2 activity is increased in patients with type 1 diabetes and vascular complications. Journal of hypertension 30: 375–383

26. Srinivasan, M., S. L. Zunt and L. I. Goldblatt, 2020 Oral epithelial expression of angiotensin converting enzyme-2: Implications for COVID-19 diagnosis and prognosis

27. Steyaert, S., G. Trooskens, J. R. Delanghe and W. Van Criekinge, 2020Differential methylation as a mediator of COVID-19 susceptibility. BioRxiv

28. Theda, C., S. H. Hwang, A. Czajko, Y. J. Loke, P. Leong et al., 2018Quantitation of the cellular content of saliva and buccal swab samples. Scientific reports 8: 1–8

29. Wysocki, J., L. Garcia-Halpin, M. Ye, C. Maier, K. Sowers et al., 2013Regulation of urinary ACE2 in diabetic mice. American Journal of Physiology-Renal Physiology 305: F600–F611

30. Xu, R., B. Cui, X. Duan, P. Zhang, X. Zhou et al., 2020Saliva: potential diagnostic value and transmission of 2019-nCoV. International Journal of Oral Science 12: 1–6

31. Xudong, X., C. Junzhu, W. Xingxiang, Z. Furong and L. Yanrong, 2006 Age-and gender-related difference of ACE2 expression in rat lung. Life sciences 78: 2166–2171

32. Zhu, N., D. Zhang, W. Wang, X. Li, B. Yang et al., 2020China Novel Coronavirus Investigating and Research Team. A novel coronavirus from patients with pneumonia in China, 2019. N Engl J Med 382: 727–733

33. Zill, P., T. C. Baghai, C. Schüle, C. Born, C. Früstück et al., 2012DNA methylation analysis of the angiotensin converting enzyme (ACE) gene in major depression. PLoS One 7: e40479

